# Development and Validation of the Patient History COVID-19 (PH-Covid19) Scoring System: A Multivariable Prediction Model of Death in Mexican Patients with COVID-19

**DOI:** 10.1101/2020.09.05.20189142

**Authors:** J. Mancilla-Galindo, J. M. Vera-Zertuche, A. R. Navarro-Cruz, O. Segura-Badilla, G. Reyes-Velázquez, F. J. Tepepa-López, P. Aguilar-Alonso, J. de J. Vidal-Mayo, A. Kammar-García

## Abstract

We sought to develop and validate a multivariable prediction model of death in Mexican patients with COVID-19, by using demographic and patient history predictors. We conducted a national retrospective cohort in two different sets of patients from the Mexican COVID-19 Epidemiologic Surveillance Study. To develop the model, we included 264,026 patients tested for SARS-CoV-2 between February 28 and May 30, 2020. To validate the model, 592,160 patients studied between June 1 and July 23, 2020 were included. Patients with a positive RT-PCR for SARS-CoV-2 and complete unduplicated data were eligible. Demographic and patient history variables were analyzed through Multivariable Cox regression models to evaluate predictors to be included in the prognostic scoring system called PH-Covid19. 83,779 patients were included to develop the model; 100,000, to validate the model. Eight predictors (age, sex, diabetes, COPD, immunosuppression, hypertension, obesity, and CKD) were included in the PH-Covid19 scoring system (range of values: −2 to 25 points). The predictive model has a discrimination of death of 0.8 (95%CI:0.796-0.804). The PH-Covid19 scoring system was developed and validated in Mexican patients to aid clinicians to stratify patients with COVID-19 at risk of fatal outcomes, allowing for better and efficient use of resources.

## Introduction

The coronavirus disease (COVID-19) pandemic is the global phenomenon which is shaping modern societies in the year 2020, a reason why the severe acute respiratory coronavirus 2 (SARS-CoV-2) has been named the once-in-a-century pathogen that scientists and global leaders had been worrying for [1]. Demographic and patient history risk factors for fatal outcomes in patients with COVID-19 have been characterized in large national cohorts [2–5], and broadly include old age, sex (men), comorbidities, deprivation (a correlate of poverty), and belonging to certain ethnic groups. Other clinical, radiological, and laboratory parameters at presentation have also been studied as risk factors for disease progression and death [5,6].

Low and middle-income countries often suffer from inadequate healthcare due to the lack of equipment, poor organization, and scarce qualified healthcare professionals. Thus, what works in high-income countries may not work in low-income countries [7]. Mexico has been one of the most affected countries by COVID-19, and disparate differences in patient outcomes have been noted be related to inequalities in healthcare access. Therefore, there is a pressing need to develop accessible and simple tools to aid clinicians providing medical attention in the most unfavored regions of Mexico.

Several diagnostic and prognostic models have been developed to be used in patients with COVID-19 [8]. However, most models include laboratory and radiographic variables which would be nearly impossible to collect in low-resource settings. Furthermore, these models have seldom been validated, are often inadequately reported, or are overfitted due to a large predictor to outcome ratio [8–10], reasons that may limit their usefulness in real-world settings. Developing and validating models that only require demographic and patient history data, by using large national or multinational cohorts, may be a way to overcome these shortcomings to provide useful tools to clinicians in low-resource regions.

Therefore, we sought to develop and validate a multivariable prediction model of death in Mexican patients with COVID-19, by using demographic and patient history predictors.

## Method

### Study Design

We conducted a national retrospective cohort in two different sets of patients from the Mexican COVID-19 Epidemiologic Surveillance Study [11] to develop and validate a multivariable prediction model of death in Mexican patients with COVID-19. Patient history variables were used as predictors of death as the outcome of interest. Blind assessment was not required since these are objective variables unlikely subjected to bias.

To develop the model, we included 264,026 patients studied between 28 February and 30 May, 2020. All patients with a positive RT-PCR for SARS-CoV-2 were included to maximize the power and generalizability of results. Patients with incomplete data were excluded, whereas patients with the same demographic, clinical, and follow-up variables were considered duplicated and only one entry was kept.

To validate the model, we included 592,160 patients studied between 1 June and 23 July, 2020. Only patients with a positive RT-PCR for SARS-CoV-2 and complete unduplicated data were included to validate the model. We further performed simple random sampling of positive cases to increase statistical power in approximately 15% with respect to the sample used for developing the model.

### Source of Data

Data is collected and regularly updated by the Mexican Secretariat of Health and is available in the Open Data platform of the Federal Government of Mexico [12]. A historical repository of individual datasets starting on 12 April 2020 is available through the General Directorate of Epidemiology [13]. Patients who met criteria of suspected COVID-19 case and were subsequently tested for SARS-CoV-2 were included in the study, starting on late February 2020 when the first suspected cases arrived in Mexico. Two diagnostic strategies are outlined in the National COVID-19 Epidemiologic Surveillance Plan [11]: 1. testing of 10% of ambulatory patients with mild symptoms of respiratory disease and 100% of patients with respiratory distress at evaluation in one of the 475 monitoring units of viral respiratory disease (USMER, for its acronym in Spanish) which are strategically distributed to be representative of the Mexican population, and 2. testing 100% of patients who meet diagnostic criteria of Severe Acute Respiratory Infection (defined as shortness of breath, temperature ≥38 °C, cough, and ≥1 of the following: chest pain, tachypnea, or acute respiratory distress syndrome) who seek medical attention in non-USMER units.

Healthcare professionals collecting a diagnostic specimen are required to fill out a format containing demographic and patient history variables. Follow-up of all suspected COVID-19 cases is registered by accredited hospital epidemiologists (inpatients) and the responsible healthcare professional of every Local Health Jurisdiction (ambulatory patients), who ultimately upload data into the Respiratory Diseases Epidemiologic Surveillance System. Results of diagnostic RT-PCR for SARS-CoV-2 are directly uploaded by the diagnostic facility; accreditation of diagnostic procedures by the Mexican Institute of Diagnostics and Epidemiological Reference is required to upload results. Reporting of deaths is obligatory and must be done in less than 48 hours after occurrence. One caveat to this reporting method is that patients who are tested more than once in different jurisdictions may be duplicated. No variables that could lead to identification of patients are provided in datasets. Thus, the only way to eliminate duplications is through matching of cases with equal demographic and clinical variables. Specific information of treatments is not released.

Variables provided in the datasets are: origin (USMER, non-USMER), healthcare provider, state, birthplace, place of residency, nationality, indigenous language speaker, migratory status, type of medical attention (hospitalization/ambulatory), admission date, symptom onset date, invasive mechanical ventilation (intubation [yes/no]), admission to ICU (yes/no), pneumonia (yes/no), date of death, contact with confirmed COVID-19 cases (yes/no), SARS-CoV-2 RT-PCR result (positive, negative, or pending), age, sex, current pregnancy, and the following comorbidities (yes/no): diabetes, hypertension, obesity, cardiovascular disease (CVD), chronic kidney disease (CKD), immunosuppression, asthma, chronic obstructive pulmonary disease (COPD), and smoking.

### Statistical Analysis

Descriptive data were calculated and are provided as frequencies, percentages, or mean with standard deviation (SD). Characteristics of patients in the model development and validation cohorts were compared through Student’s t-test or X^2^ A Cox regression model was applied to predict the risk of death. The risk of death was assessed through univariate analysis of the following variables: age, sex, current pregnancy, diabetes, COPD, asthma, immunosuppression, hypertension, CVD, obesity, CKD, smoking, and time from symptom onset to medical attention. Risk factors associated with death which were statistically significant (p< 0.05) were included to develop the multivariate regression model. All variables with a level of significance p< 0.1 were considered in the Cox regression model by using the Enter method. Variables that kept a level of significance p< 0.05 were used in the final model, which was evaluated through Harrell’s C-statistic to determine its discrimination of death.

A scoring system was developed in accordance to the model proposed by Sullivan LM, et al. [14] Each risk factor was organized into categories and the reference value was determined as follows. Age was entered into the Cox regression model as a continuous variable and further categorized into 10 sets of years (< 20, 20-29, 30-39, 40-49, 50-59, 60-69, 70-79, 80-89, 90-99, and > 90); the midpoint between the nine values of each category was set as the reference value (*W_ij_*). The reference for age as a risk factor (W_iREF_) was set in the 20-29 years category since this group had the lowest mortality in a previous study performed in Mexican patients with COVID-19 [3]. The rest of the risk factors were modeled as dummy variables, setting the absence of comorbidities and being woman (sex) as the reference values. Each W_iREF_ was subtracted from *W_ij_* and multiplied by the regression coefficient (β_*i*_) of the risk factor to determine units of regression of distancing for every risk factor in the reference category β_i_(W_ij_ - W_iREF_). A constant B, defined as the constant increase in units of risk for each 5-year increase, was obtained by multiplying β_*i*_ • 5. Values for B were 0.25 in this study. Scores for each category were obtained with the equation β_i_(W_ij_ – W_iREF_)B. For every point in the risk score, the estimated risk of death (p) was calculated with the Cox proportional hazards regression analysis:

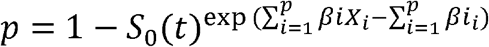

Where: *S_0_(t)* is the average survival according to mean values of every risk factor; 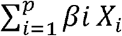 is substituted with each value of the risk score, times the B constant, plus the reference age value according to the β_*i*_ of age; and 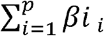 is the sum of every β_*i*_ times the proportion or mean value of every risk factor.

To validate the model, we calculated the risk score for every patient and applied the Cox proportional hazards regression analysis. Estimated risks were obtained from both the scoring system and the observed risk in the regression analysis. The values obtained for patients in the validation cohort were distributed in percentiles (1 to 99) to determine the punctuation categories. The estimated and observed risks were compared in each category of punctuation (percentiles 25, 50, 75, and 99).

The Kaplan-Meier analysis was performed to determine survival in each category of the scoring system, and a Cox regression analysis, to determine increases in the lethality risk for each category.

The association between the risk score and the probability of other adverse events (hospitalization, invasive mechanical ventilation, pneumonia, and admission to ICU) were also studied. The frequencies of each adverse event for every category in the scoring system were quantified and a binomial logistic regression analysis was performed to determine the risk of each adverse event according to the risk score; logarithms of the odds ratios (OR) were graphed to stablish the scoring value at which risk for every adverse event is increased.

All statistical analyses were performed using the SPSS software v.21 and the R statistical software v.3.6.2; figures were created in GraphPad Prism v.6. A value of p< 0.05 was used to stablish statistical significance.

## Results

Out of 264,026 patients in the model development cohort, 84,627 had a positive RT-PCR for SARS-CoV-2, 140,553 were negative, and 38,846 had pending results. After exclusion of patients with incomplete data and duplicated registries, 83,779 patients with a positive test were included to develop the model. Among the 592,160 patients in the model validation cohort, 256,488 patients had a positive result, 253,447 were negative, and 82,225 had unreported results. After excluding duplicated and incomplete registries, and random sampling of positive cases, 100,000 patients were included to validate the model. Descriptive values of demographic characteristics, patient history, and outcomes of patients (survivors and non survivors) in both cohorts are provided in Table 1.

**Table 1.** Demographic characteristics, patient history data, and outcomes in the model development and validation cohorts.

|  | Development cohort,<br>positive cases |  |  | Validation cohort,<br>positive cases |  |  |
| --- | --- | --- | --- | --- | --- | --- |
|  | Total<br>n=83,779 | Survivors<br>n=74,551 | Non-survivors<br>n=9,228 | Total<br>n=100,000 | Survivors<br>n=94,722 | Non-survivors<br>n=5,278 |
| Sex |  |  |  |  |  |  |
| Women, n (%) | 36,393 (43.4) | 33,370 (44.8) | 3,023 (32.8)<sup>a</sup> | 48,129 (48.1) | 46,304 (48.9) | 1,825 (34.6)<sup>a</sup> |
| Men, n (%) | 47,386 (56.6) | 41,181 (55.2) | 6,205 (67.2)<sup>a</sup> | 51,871 (51.9) | 48,418 (51.1) | 3,453 (65.4)<sup>a</sup> |
| Age, years | 46.3 (15.9) | 44.6 (15.4) | 60 (14.2)<sup>a</sup> | 43.9 (16.5) | 42.9 (16.1) | 61.3 (14.5)<sup>a</sup> |
| Smoking, n (%) | 6,966 (8.3) | 6,092 (8.2) | 874 (9.5)<sup>a</sup> | 6,935 (6.9) | 6,538 (6.9) | 397 (7.5) |
| Pregnancy, n (%) | 537 (0.6) | 523 (0.7) | 14 (0.2)<sup>a</sup> | 729 (0.7) | 724 (0.8) | 5 (0.1)<sup>a</sup> |
| Contact with confirmed case,<br>n (%) | 26,408 (31.3) | 25,127 (33.7) | 1,100 (11.9)<sup>a</sup> | 52,047 (52) | 50,563 (53.4) | 1,484 (28.1)<sup>a</sup> |
| Comorbidities, n (%) |  |  |  |  |  |  |
| Diabetes | 14,735 (17.6) | 11,235 (15.1) | 3,500 (37.9)<sup>a</sup> | 14,036 (14) | 12,010 (12.7) | 2,026 (38.4)<sup>a</sup> |
| COPD | 1,695 (2) | 1,183 (1.6) | 512 (5.5)<sup>a</sup> | 1,244 (1.2) | 1,020 (1.1) | 224 (4.2)<sup>a</sup> |
| Asthma | 2,466 (2.9) | 2,258 (3) | 208 (2.3)<sup>a</sup> | 2,543 (2.5) | 2,427 (2.6) | 116 (2.2) |
| Immunosuppression | 1,300 (1.6) | 1,020 (1.4) | 280 (3)<sup>a</sup> | 973 (1) | 848 (0.9) | 125 (2.4)<sup>a</sup> |
| Hypertension | 17,609 (21) | 13,696 (18.4) | 3,913 (42.4)<sup>a</sup> | 17,162 (17.2) | 14,970 (15.8) | 2,192 (41.5)<sup>a</sup> |
| CVD | 2,178 (2.6) | 1,640 (2.2) | 538 (5.8)<sup>a</sup> | 1,875 (1.9) | 1,601 (1.7) | 274 (5.2)<sup>a</sup> |
| Obesity | 17,244 (20.6) | 14,752 (19.8) | 2,492 (27)<sup>a</sup> | 18,161 (18.2) | 16,698 (17.6) | 1,463 (27.7)<sup>a</sup> |
| CKD | 1,944 (2.3) | 1,293 (1.7) | 651 (7.1)<sup>a</sup> | 1,401 (1.4) | 1,131 (1.2) | 270 (5.1)<sup>a</sup> |
| Time from symptom onset to<br>medical attention, days | 4.3 (3.2) | 4.3 (3.2) | 4.5 (3.1)<sup>a</sup> | 4.3 (3.1) | 4.2 (3.1) | 5.1 (3.2)<sup>a</sup> |
| Hospitalization, n (%) | 29,535 (35.3) | 21,257 (28.5) | 8,278 (89.7)<sup>a</sup> | 19,316 (19.3) | 14,753 (15.6) | 4,563 (86.5)<sup>a</sup> |
| Intubation, n (%) | 2,923 (3.5) | 953 (1.3) | 1,970 (21.3)<sup>a</sup> | 2,260 (2.3) | 778 (0.8) | 1,482 (28.1)<sup>a</sup> |
| Pneumonia, n (%) | 22,785 (27.2) | 15,644 (21) | 7,141 (77.4)<sup>a</sup> | 16,742 (16.7) | 12,122 (12.8) | 4,620 (87.5)<sup>a</sup> |
| ICU admission, n (%) | 2,791 (3.3) | 1,359 (1.8) | 1,432 (15.5)<sup>a</sup> | 2,383 (2.4) | 1,419 (1.5) | 964 (18.3)<sup>a</sup> |
| Case-Fatality rate (%) | 11.02% | - | - | 5.3% | - | - |

Data are presented as mean values (SD), unless otherwise specified.
a: Statistical significance regarding Survivors

Variables included in the Cox regression model are presented in Table 2. Eight risk factors were included in the model; the only quantitative variable was age. Age, diabetes, and CKD were associated with the greatest increases in death. The predictive model has a discrimination of 0.8 (95%CI:0.796-0.804) and an average survival of 0.903 with the mean values for every risk factor. The Patient History COVID-19 (PH-Covid19) scoring system assigns a score to every risk factor ultimately included in the predictive model (Table 3); the sum of scores for all risk factors included ranges from −2 to 25 points. Predicted probabilities of death in patients with a positive test for SARS-CoV-2 for every possible total value in the scoring system range from 0.74% to 99.82% (Table 4).

**Table 2.** Risk factors associated with death in Mexican patients with a positive diagnostic test for SARS-CoV-2 (model development cohort)

|  | Regression coefficient | Standard error | HR (95%CI) | p value | Mean or proportion |
| --- | --- | --- | --- | --- | --- |
| Age | 0.050 | 0.001 | 1.05 (1.05-1.05) | <0.0001 | 46.27 |
| Sex (men) | 0.473 | 0.022 | 1.6 (1.54-1.68) | <0.0001 | 0.57 |
| Diabetes | 0.437 | 0.023 | 1.55 (1.48-1.62) | <0.0001 | 0.18 |
| COPD | 0.136 | 0.047 | 1.15 (1.05-1.26) | 0.004 | 0.02 |
| Immunosuppression | 0.300 | 0.062 | 1.35 (1.19-1.52) | <0.0001 | 0.02 |
| Hypertension | 0.206 | 0.024 | 1.23 (1.17-1.29) | <0.0001 | 0.21 |
| Obesity | 0.337 | 0.024 | 1.4 (1.34-1.47) | <0.0001 | 0.21 |
| CKD | 0.620 | 0.042 | 1.86 (1.71-2.02) | <0.0001 | 0.02 |

HR: Hazard ratio, 95%CI: 95% Confidence Interval.
COPD: Chronic obstructive pulmonary disease, CKD: Chronic kidney disease

**Table 3.** The PH-Covid19 risk score to predict death in patients with COVID-19

| Risk Factor | Score |
| --- | --- |
| Age |  |
| <20 | -2 |
| 20-29 | 0 |
| 30-39 | 2 |
| 40-49 | 4 |
| 50-59 | 6 |
| 60-69 | 8 |
| 70-79 | 10 |
| 80-89 | 12 |
| 90-99 | 14 |
| >99 | 15 |
| Sex |  |
| Male | 2 |
| Comorbidities |  |
| Diabetes | 2 |
| COPD | 1 |
| Immunosuppression | 1 |
| Hypertension | 1 |
| Obesity | 1 |
| CKD | 2 |

Range of values: -2 a 25
COPD: Chronic obstructive pulmonary disease, CKD: Chronic kidney disease.

**Table 4.** Estimated risk of death according to every possible score in the PHCovid19 score, in Mexican patients with a positive test for SARS-CoV-2

| **Total value** | **Estimated Risk** | **Total value** | **Estimated Risk** |
| --- | --- | --- | --- |
| -2 | 0.74% | 12 | 21.72% |
| -1 | 0.94% | 13 | 26.98% |
| 0 | 1.21% | 14 | 33.22% |
| 1 | 1.55% | 15 | 40.45% |
| 2 | 1.99% | 16 | 48.60% |
| 3 | 2.55% | 17 | 57.46% |
| 4 | 3.26% | 18 | 66.63% |
| 5 | 4.17% | 19 | 75.56% |
| 6 | 5.32% | 20 | 83.62% |
| 7 | 6.77% | 21 | 90.20% |
| 8 | 8.61% | 22 | 94.94% |
| 9 | 10.92% | 23 | 97.83% |
| 10 | 13.80% | 24 | 99.27% |
| 11 | 17.36% | 25 | 99.82% |

Baseline characteristics and outcomes of patients in the validation cohort were statistically significantly different from those in the model development cohort, except for time from symptom onset to medical attention (Table 1). Results of the Cox regression model applied to the validation cohort are provided in Supplementary Table S1. Calculated scores in the validation cohort reflected the following distribution according to percentiles: −2 to 2 points, percentile 1 to 25; 3 to 5 points, percentile 25-50; 6 to 8 points, percentile 51 to 75; and 9 to 15 points, percentile 76 to 99. Patients > 99 percentile were considered as extreme values. Estimated risks and the observed risks of death obtained from the Cox proportional hazards regression analysis, in relation to the scores in each percentile, are presented in Fig. 1. The estimated and observed risks were similar for each group and were strongly correlated (r = 0.98, R^2^ = 0.96, p< 0.0001).

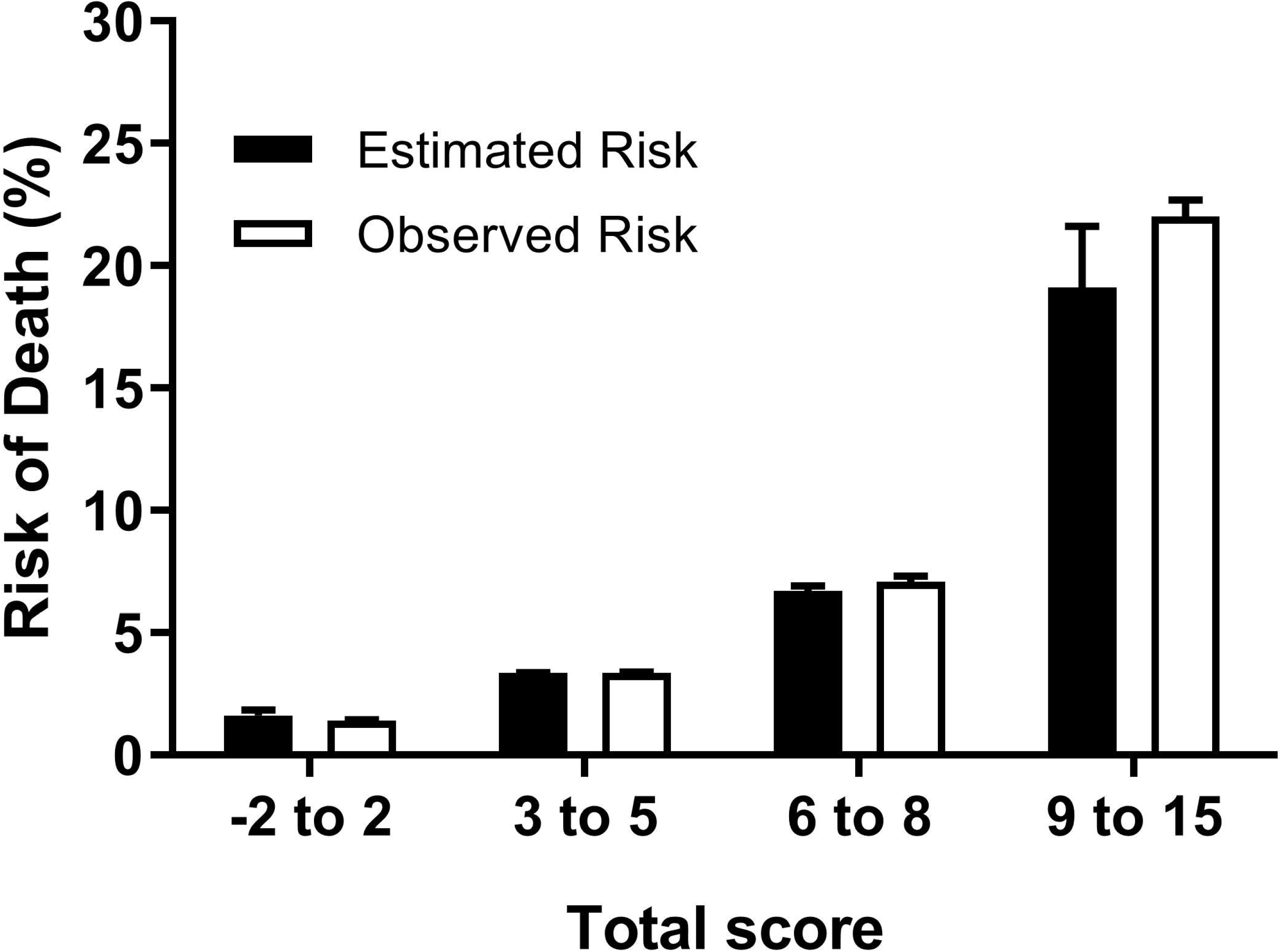

In the survival comparison between categories generated according to percentiles (Fig. 2), survival was decreased with increasing risk scores (−2 to 2 points, 99.6%; 3 to 5 points, 98.6%; 6 to 8 points; 95.7%, and 9 to 15 points, 84.3%, p< 0.0001 for all categories). A subsequent Cox regression analysis was performed to determine the risk of death for each individual category compared to the category comprising the lower percentile (−2 to 2 points): 2 to 5 points, HR:3.54 (95-CI:2.85-4.39); 6 to 8 points, HR:10.67 (95%CI:8.78-12.98); 9 to 15 points, HR:41.9 (95%CI:34.7-50.7); and > 15 points, HR:87.6, (95%CI:70.1-109.5). In accordance with this, risk groups were derived from the scoring system: Low (−2 to 2 points), Medium-Low (3 to 5 points), Medium (6 to 8 points), Medium-High (9 to 15 points), and High (>15 points).

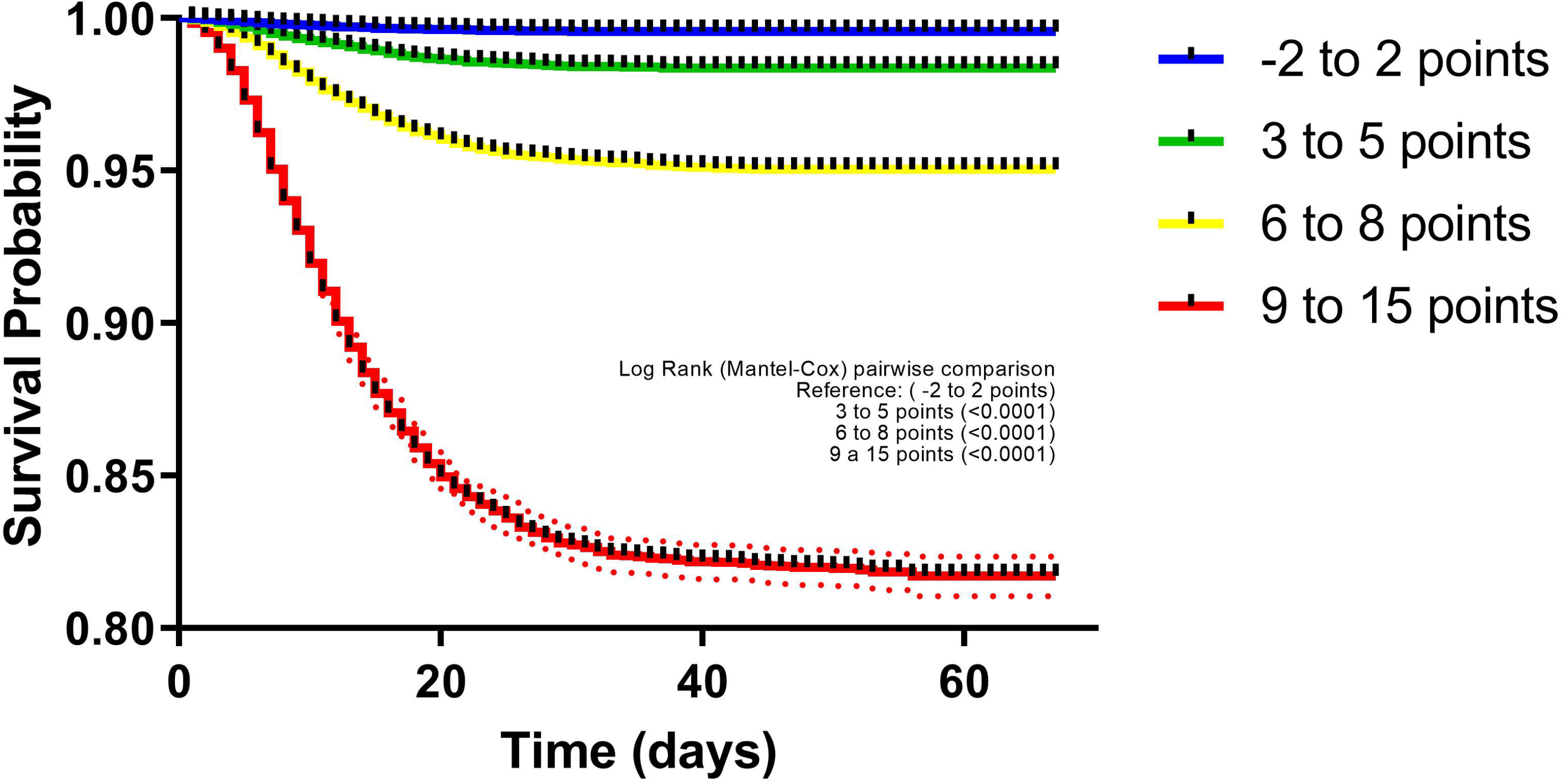

The prevalence for every adverse event in each category of the scoring system are provided in Fig. 3; results of the logistic regression analysis to determine the risk of adverse events for each risk group in the PH-Covid19 scoring system are given in Supplementary Table S2. Tendencies of risk increasement for each adverse event with augmenting scores (Supplementary Figure S1) reflect that risk for any adverse event starts at a value of 5 points.

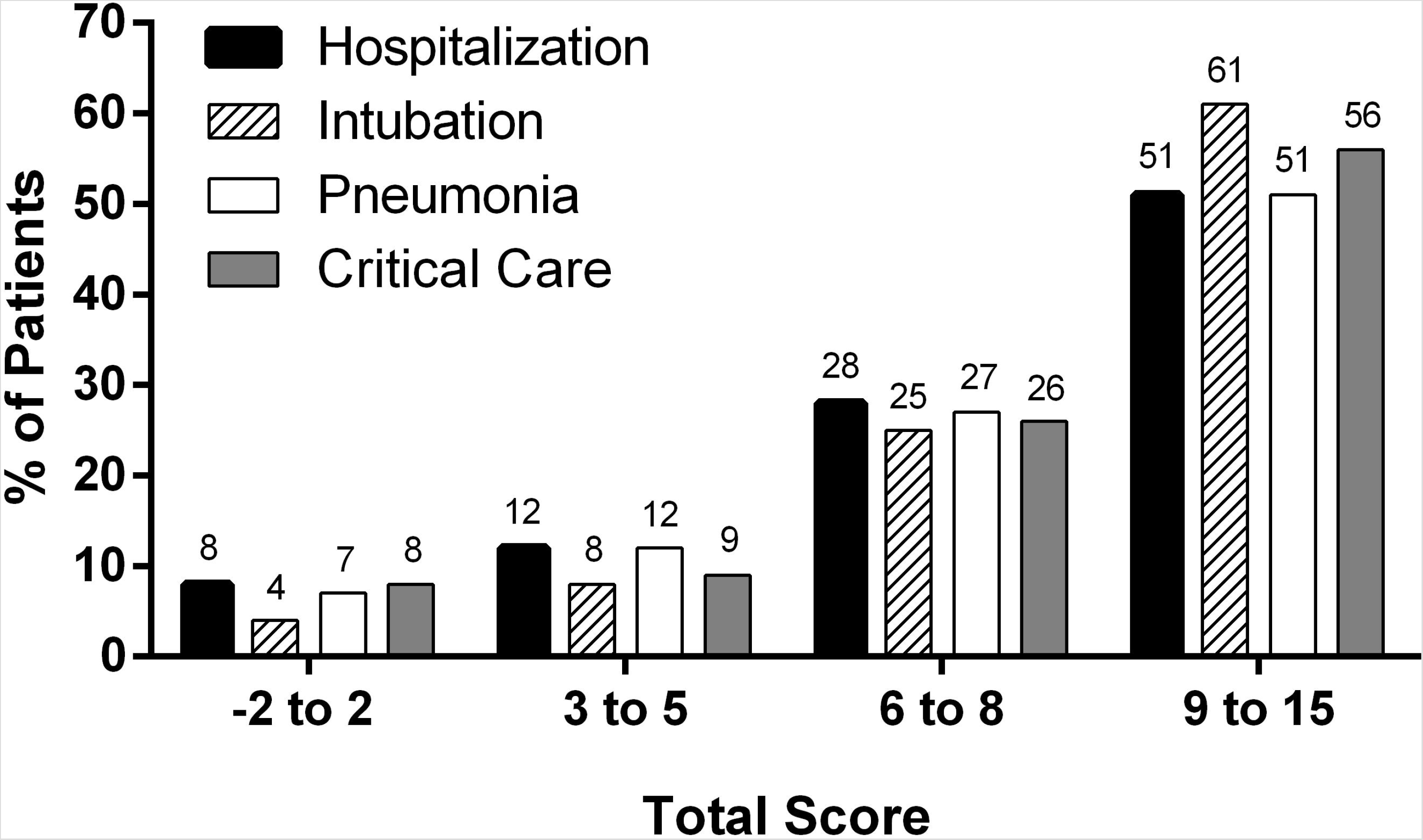

## Discussion

We have developed and validated the PH-Covid19 score, a multivariable prediction model of death in Mexican patients with COVID-19, by using different datasets from the Mexican COVID-19 Epidemiologic Surveillance Study. This scoring system has been created to aid clinicians working in resource-strained conditions to early stratify patients with COVID-19 according to their risk of fatal outcomes, without the need to perform laboratory or imaging studies.

Similar to other studies, we found that older age is the main risk factor for dying from COVID-19, with every 10-year increase associated with the largest increases in hazard ratio [2,5,15]; in the PH-Covid19 score, 10-year increases added two points, starting from 30 years, and being < 20 years subtracted 2 points. Sex (men), which increases two points in our score, was correlated with death in our study and others [2,3,5,6,15]. Diabetes and CKD resulted in two-point increases in the score. Other studies had similar findings to ours [3,4,6,15]; one study found an uncertain increased risk attributable to diabetes (HR:1.14, 95%CI:0.91-1.43) in critically-ill patients [5]; in another study, uncontrolled diabetes further increased the risk (HR:1.95, 95%CI:1.83–2.08) with respect to controlled diabetes (HR:1.31, 95%CI:1.24-1.37), while CKD stages 4-5 caused a greater increase (HR:2.52, 95%CI:2.33-2.72) than CKD stages 3a-3b (HR:2.52, 95%CI:2.33-2.72) [2]. Obesity had the third strongest association with death, resulting in a 1-point increase in our scoring system; other studies have had similar findings [3–15]. Adjusted risk of death by obesity occurred with gradual increases in higher obesity classes^2^ and no evident or clear risk was identified in another study for class I and II obesity, whereas class III obesity was significantly correlated with death[5]. This last study, however, included only patients admitted to ICUs who had a high prevalence of comorbidities, which are independent risk factors for ICU admission [3].

Hypertension was a risk factor for death similar to other studies [3,4], and resulted in a 1-point increase in the scoring system. In one large study, adjusted risk of death for hypertension adjusted by all covariates apparently reduced the risk (HR:0.89, 95%CI:0.85–0.93), likely reflecting an artificial reduction of risk driven by diabetes and obesity since age and sex adjusted risk increased the risk (HR:1.09, 95%CI:1.05–1.14)[2]. COPD also increased 1 point in our model, being also associated to death in other studies [2–4,15].

Results from our validation did not provide different patient history predictors which could enhance the performance of our model. Thus, model updating was not required. Some of our findings raise concerns regarding quality and access to healthcare in Mexico. 10.3-13.5% of patients who died in our cohorts did not receive in-hospital care at any moment of the disease, only 15.5- 18.3% of patients were admitted to an ICU before dying, and only 21.3-28.1% of patients who died were intubated; conversely, intubation in survivors was unusually low (0.8-1.3%). In other studies of hospitalized-only patients, 53-72% of non-survivors were admitted to an ICU and 51-59% received invasive mechanical ventilation [6,16,17]. Observed mortality in COVID-19 patients under 60 years is lower when access to healthcare is not a limitation [18]; non-survivors in our cohorts were younger (mean age 60-61.4 years) than those in other studies (67-80 years) [5,6,15,19].

Patients diagnosed with pneumonia in both cohorts were 12.8-21% among survivors and 77.4-87.5% of non-survivors. These numbers are low compared to the prevalence of chest CT-scan abnormalities which occur in 67.3-70.8% of asymptomatic/pre-symptomatic patients [20,21], 95.5% of patients with mild COVID-19 [22], and 98% of all COVID-19 patients included in a meta-analysis [23]. Chest X-ray, on the other hand, may be normal in up to 63% of patients with early COVID-19 pneumonia [24]. Nonetheless, the low proportion of pneumonia in non-survivors suggests non-optimal diagnosis of pneumonia may be occurring in Mexico. The lack of an operational definition may have contributed since clinicians could have defined pneumonia differently based on clinical and/or radiographical findings. Other possibilities should be explored, including knowledge of Mexican clinicians on how to diagnose pneumonia and access to radiological studies during the pandemic in low-resource settings.

Three prognostic COVID-19 models have been developed in Mexican patients. The LOW-HARM model is a 100-point scoring system calculated by inputting patient history and laboratory values, setting 65 points as the cut-off value of greater discrimination of risk of death [25]. Prediction of death (0.81, 95%CI:0.77–0.84) is similar to the PH-Covid19 scoring system (0.80, CI95%:0796-0.804), which advantageously only requires patient history predictors. Another scoring system uses age (cut-off 65 years), comorbidities, and pneumonia to predict death [26]. This model was accurate at predicting death and other adverse events but has the limitation of not accounting for the large increases in risk for every 10-year category or similar. Also, the model by Bello-Chavoya, et al. was developed using one dataset of the Mexican Epidemiologic Surveillance Study, which unfortunately had no operational definition for pneumonia as discussed earlier. A third model was developed and validated to predict the risk of admission to ICU; the ABC-GOALS was developed in three versions: clinical, clinical + laboratory, and clinical + laboratory + imaging predictors [27].

In one systematic review of existing prediction models for diagnosis and prognosis of COVID-19, the use of any of the reviewed models was discouraged since, out of 91 diagnostic and 50 prognostic models, all were at high risk of bias due to methodological constraints and poor reporting [8]. Predictive models of death (eight) often excluded patients that had not developed the outcome of interest, did not account for censorship, inadequately reported discrimination and calibration of the model, and had a high risk of bias according to PROBAST evaluation, despite authors claiming good global performances of their models. We have addressed these concerns in our development and validation of the PH-Covid19 scoring system.

Another strength of our study is that we were able to perform a type 3 analysis according to TRIPOD by using individual datasets to develop and validate our model; this design allows for external validation of the performance of a model [28]. It is worth highlighting that sample sizes in both cohorts include thousands of patients, which adds robustness to our model.

One limitation of our study is that certain diseases (cancer, hematological malignancies, and neurologic diseases) and specific states of a disease (obesity class, former or current smoker, and control of diabetes, hypertension, and asthma) which increase the risk of dying from COVID-19 [2] were not studied since they are not provided in the datasets. However, our model accounts for the main risk factors associated with death in patients with COVID-19, and not requiring inputting specific disease states makes it easier to be used by clinicians while minimizing the risk of not having enough data to use the score with precision.

Another limitation is that the epidemiologic surveillance strategy in Mexico allows testing of only 10% of ambulatory patients. Furthermore, the operational definition of suspected COVID-19 case used in Mexico until August 24, 2020 had a low sensitivity (58.2%), but a high specificity (63.7%) compared to that used by the CDC (85.8% and 25.8%, respectively) [29]. Altogether, this means that our cohorts may include very few patients with asymptomatic COVID-19 and less patients with mild COVID-19 compared to other national datasets with higher testing rates. Since it was not possible to determine the exact number of patients with mild disease in our cohorts, we can only indirectly suggest that mild-disease patients could comprise around 71.5 and 84.4% of patients according to the fraction of non-hospitalized patients who survived, a number high enough to permit the use of this score in patients with mild-to-severe COVID-19.

The PH-Covid19 score was created to be used in limited-resource settings where access to laboratory and imaging studies may be restricted. In places where this is not a limitation and for patients who are likely to have already been admitted to hospital (critical patients), other prognostic models may have a better performance than ours. However, clinicians should consider that most models have not been validated before deciding to use any COVID-19 diagnostic or prognostic model.

The PH-Covid19 score uses patient history predictors which are frequently known at the first contact with a patient, or can be interrogated rapidly, to predict death in patients with COVID-19. This score was developed and validated in Mexican patients to be used in low-resource settings where obtaining laboratory and radiographic studies may not be immediately possible. This score will aid clinicians to stratify patients with COVID-19 at risk of fatal outcomes to use healthcare resources more efficiently.

## Data Availability

all data is free for your consultation

https://www.gob.mx/salud/documentos/datos-abierto-bases-historicas-direccion-general-de-epidemiologia

## Ethical disclosures

This article is a retrospective study of an open-source dataset of patients in Mexico. The Ministry of Health of Mexico approved the recollection of information.

## Acknowledgements

J M-G would like to thank “Dirección General de Calidad y Educación en Salud” for supporting his participation in “Programa Nacional de Servicio Social en Investigación en Salud”.

## References

1. Gates B. Responding to Covid-19 — A Once-in-a-Century Pandemic? New England Journal of Medicine 2020; 382: 1677–1679.

2. Williamson EJ, et al Factors associated with COVID-19-related death using OpenSAFELY. Nature Springer US, 2020; 584: 430–436.

3. Kammar-García A, et al Impact of Comorbidities in Mexican SARS-CoV-2-Positive Patients: A Retrospective Analysis in a National Cohort. Revista de investigacion Clinica 2020; 72: 151–8.

4. Guan W, et al Comorbidity and its impact on 1590 patients with COVID-19 in China: a nationwide analysis. European Respiratory Journal 2020; 55: 2000547.

5. Gupta S, et al Factors Associated With Death in Critically Ill Patients With Coronavirus Disease 2019 in the US. JAMA Internal Medicine 2020; 02115: 1–11.

6. Zhou F, et al Clinical course and risk factors for mortality of adult inpatients with COVID-19 in Wuhan, China: a retrospective cohort study. The Lancet 2020; 395: 1054–1062.

7. Roder-DeWan S. Health system quality in the time of COVID-19. The Lancet Global Health 2020; 8: e738–e739.

8. Wynants L, et al Prediction models for diagnosis and prognosis of covid-19: Systematic review and critical appraisal. The BMJ 2020; 369: m1328.

9. Collins GS, van Smeden M, Riley RD. COVID-19 prediction models should adhere to methodological and reporting standards. European Respiratory Journal. Published online: 23 July 2020. doi: 10.1183/13993003.02643-2020.

10. Hooli S, King C. Generalizability of Coronavirus Disease 2019 (COVID-19) Clinical Prediction Models. Clinical Infectious Diseases 2020; 71: 897–897.

11. Directorate General of Epidemiology MEX. Standardized Guideline for Epidemiologic and Laboratory Surveillance of viral respiratory diseases, May 2020 [Spanish]. Mexican Secreatariat of Health. Mexico City, 2020, p. 1–80 (https://www.gob.mx/cms/uploads/attachment/file/552972/Lineamiento_VE_y_Lab_Enf_Viral_20.05.20.pdf). Accessed 17 August 2020.

12. Health Ministry MEX. Information regarding COVID-19 cases in Mexico [Spanish]. Datos Abiertos 2020 (https://datos.gob.mx/busca/dataset/informacion-referente-a-casos-covid-19-en-mexico). Accessed 17 August 2020.

13. Directorate General of Epidemiology MEX. Historical COVID-19 Datasets [Spanish]. Datos Abiertos - Bases Históricas. 2020(https://www.gob.mx/salud/documentos/datos-abiertos-bases-histori-casdireccion-general-de-epidemiologia). Accessed 17 August 2020.

14. Sullivan LM, Massaro JM, D’Agostino RB. Presentation of multivariate data for clinical use: The Framingham Study risk score functions. Statistics in Medicine 2004; 23: 1631–1660.

15. Docherty AB, et al Features of 20 133 UK patients in hospital with covid-19 using the ISARIC WHO Clinical Characterisation Protocol: prospective observational cohort study. BMJ 2020; 369: m1985.

16. Richardson S, et al Presenting Characteristics, Comorbidities, and Outcomes Among 5700 Patients Hospitalized With COVID-19 in the New York City Area. JAMA 2020; 323: 2052.

17. Yang X, et al Clinical course and outcomes of critically ill patients with SARS-CoV-2 pneumonia in Wuhan, China: a single-centered, retrospective, observational study. The Lancet Respiratory Medicine 2020; 8: 475–481.

18. Karagiannidis C, et al Case characteristics, resource use, and outcomes of 10 021 patients with COVID-19 admitted to 920 German hospitals: an observational study. The Lancet Respiratory Medicine 2020; 2600: 1–10.

19. Wu C, et al Risk Factors Associated With Acute Respiratory Distress Syndrome and Death in Patients With Coronavirus Disease 2019 Pneumonia in Wuhan, China. JAMA Internal Medicine 2020; 180: 934.

20. Hu Z, et al Clinical characteristics of 24 asymptomatic infections with COVID-19 screened among close contacts in Nanjing, China. Science China Life Sciences 2020; 63: 706–711.

21. Wang Y, et al Clinical Outcomes in 55 Patients With Severe Acute Respiratory Syndrome Coronavirus 2 Who Were Asymptomatic at Hospital Admission in Shenzhen, China. The Journal of Infectious Diseases 2020; 221: 1770–1774.

22. Liang T, et al Evolution of CT findings in patients with mild COVID-19 pneumonia. European Radiology European Radiology, 2020; 30: 4865–4873.

23. Awulachew E, et al Computed Tomography (CT) Imaging Features of Patients with COVID-19: Systematic Review and Meta-Analysis. Radiology Research and Practice 2020; 2020: 1023506.

24. Cleverley J, Piper J, Jones MM. The role of chest radiography in confirming covid-19 pneumonia. The BMJ 2020; 370: 1–9.

25. Soto-Mota A, et al The LOW-HARM Score for Predicting Mortality in Patients Diagnosed with COVID-19: A Multicentric Validation Study. medRxiv. Published online: 5 June 2020. doi:10.1101/2020.05.26.20111120.

26. Bello-Chavolla OY, et al Predicting Mortality Due to SARS-CoV-2: A Mechanistic Score Relating Obesity and Diabetes to COVID-19 Outcomes in Mexico. The Journal of Clinical Endocrinology & Metabolism 2020; 105: 1–10.

27. Mejia-Vilet JM, et al A Risk Score to Predict Admission to Intensive Care Unit in Patients With COVID-19: The ABC-GOALS Score. medRxiv. Published online: 30 May 2020. doi: 10.1101/2020.05.12.20099416.

28. Moons KGM, et al Transparent Reporting of a multivariable prediction model for Individual Prognosis Or Diagnosis (TRIPOD): Explanation and Elaboration. Annals of Internal Medicine 2015; 162: W1–W73.

29. Instituto Mexicano del Seguro Social IMSS. Operational definition of suspected case of respiratory viral disease, including COVID-19 [Spanish]. Mexico City: Instituto Mexicano del Seguro Social (IMSS), 2020, p. 6.

